# Analyzing morphological alternations of vessels in multiple Sclerosis using SLO images of the eyes

**DOI:** 10.1101/2023.12.12.23299846

**Authors:** Asieh Soltanipour, Roya Arian, Ali Aghababaei, Raheleh Kafieh, Fereshteh Ashtari

**Author notes:** These authors contributed equally (Asieh Soltanipour, Roya Arian).

## Abstract

**Background:** Our purpose was to investigate the most relevant and discriminating clinical feature set of Scanning laser ophthalmoscopy (SLO) images, which could differentiate multiple sclerosis (MS) and healthy control (HC) patients.

**Methods:** In this work, SLO images were used for the first time to measure the most valuable manual and clinical features from some retinal structures, optic disc, cup and blood vessels, for MS and HC classifications. For this, first an age-matching algorithm along with a subject-wise k-fold cross-validation data splitting approach were applied for construction of training, validation and test dataset, minimizing the risk of model overestimation. Then, it was needed to segment the retinal structures from the SLO images, and due to the lack of ground truth for our SLO images, we took advantage of a previously proposed deep learning algorithm for anatomical segmentation using color fundus images. But owing to different imaging modalities of SLO images, we also used two stages of pre-processing and post-processing to obtain accurate results for the segmentation step. Following that, a set of manual and clinical features was measured from the segmented optic disc, cup and vessels to gain a better comprehension of the features playing an important role in classification of MS and HC images. Finally, three simple machine learning models were applied to evaluate the measured features and the most valuable and effective features were computed.

**Results:** The measured feature set from the segmented optic disc, cup and blood vessels resulted in a mean accuracy (ACC) of 83%, sensitivity (SE) of 79%, specificity (SP) of 85%, and AUROC of 84%, when testing on validation data by using a XGBoost classifier model. Furthermore, horizontally disc location, fractal dimension and intensity variation of blood vessels were selected as the most important and effective features for MS and HC classification.

**Conclusion:** The location of optic disc, fractal dimension and vessel intensity, the ratio between intensity of vessels to intensity of he whole SLO image, were selected as three most valuable features for MS and HC classification. Regarding the optic disc location, we found out the used SLO images had been captured with two different imaging techniques. So, this feature could not be trusted as the most important feature. Two other features were confirmed by one expert as clinically distinguishing features for MS and HC classification.

## 1. Introduction

Multiple sclerosis (MS) is an immune-mediated disease of the central nervous system (CNS) characterized by chronic inflammation, demyelination, gliosis, and axonal degeneration [1]. MS most frequently affects young adults, with about 2.3 million people suffering from it worldwide [1]. The disease presents with a variety of signs and symptoms, including limb weakness and paresthesia, autonomic nervous system dysfunction, and visual impairment [2]. To date, numerous structural and functional changes have been reported in the retinas of MS patients, even in the absence of any history of optic neuritis [3]. A large number of optical coherence tomography (OCT) studies have revealed that the thickness of retinal nerve fiber layer (RNFL) and ganglionic cell-inner plexiform layer (GCIPL) become thinner in MS, compared to healthy control (HC) individuals [4]; this has also been observed in other neurodegenerative conditions like Alzheimer’s disease (AD) [5] and Parkinson’s disease (PD) [6]. Indeed, the retina, as a unique window to study the brain pathology, may contain potential markers for diagnosing neurodegenerative diseases including MS, without the need to employ current invasive, costly, and time-consuming diagnostic procedures like magnetic resonance imaging (MRI) and lumbar puncture [3].

In addition to OCT, other retinal imaging modalities have also been employed to study MS. For instance, fundus camera photography has revealed optic nerve atrophy [7–9] and a decline in retinal vessel diameter [10, 11] in patients with MS. Disruption of oxygen metabolism by retinal tissue has also been demonstrated using retinal oximetry, which is a technology based on the conventional fundus photography [10, 12]. Similarly, OCT angiography (OCT-A) studies showed that vessel density (VD) of superior capillary plexus (SCP) within parafoveal and peripapillary regions decreases in eyes positive for optic neuritis [13–16]. Overall, the analysis of retinal changes in MS has mainly focused on OCT, with much less emphasis on OCT-A and fundus camera photographs thus far.

Infrared scanning laser ophthalmoscopy (IR-SLO) is an imaging technology often performed along with OCT to lock B-scans at a fixed position, thus enhancing image quality by mitigating the impact of eye motion-induced noise during image acquisition. Additionally, IR-SLO technology enables ophthalmologists to observe disease progression and response to treatments at longitudinal follow-up visits [17]. IR-SLO works by illuminating the retina with a laser beam in a raster pattern, and creating en-face two-dimensional images using the backscattered light passed through a confocal aperture [17]. The resulting images are very similar to conventional fundus photographs but have superior resolution; this is why IR-SLO is also known as monochromatic fundus photography. However, no prior research has delved into IR-SLO images in patients with MS, leading to a lack of evidence to ascertain whether any potential biomarker of MS can be found in such images.

In the current study, our aim is to investigate whether IR-SLO images exhibit distinctive features specific for MS, thereby enabling differentiation between MS patients and HC individuals. We first applied a number of image processing algorithms to extract 26 features concerning the optic disc and retinal vessels; notably, certain features, such as vessel tortuosity and fractal dimension, have not been previously investigated in an MS population previously, even when considering studies on fundus camera photographs. Subsequently, the discriminating capacity of each feature was quantitively calculated using well-known feature importance techniques. This study would be the first comprehensive analysis of IR-SLO images in patients with MS.

## 2. Methods and Materials

In this study, our goal was to evaluate the fundamental clinical characteristics that effectively differentiate SLO images of MS and HC. To accomplish this objective, we employed a classification application. Initially, we generated a set of manual features derived from anatomically segmented structures, including the optic disc, cup, and blood vessels. Subsequently, we assessed these features using basic machine learning (ML) classifiers and determined the value and significance of each characteristic through three distinct methods. This process aimed to pinpoint the most critical and valuable features from the entire feature set for the categorization of SLO images into MS and HC groups. A brief overview of our proposed approach is illustrated in Figure 1.

**Figure 1.**
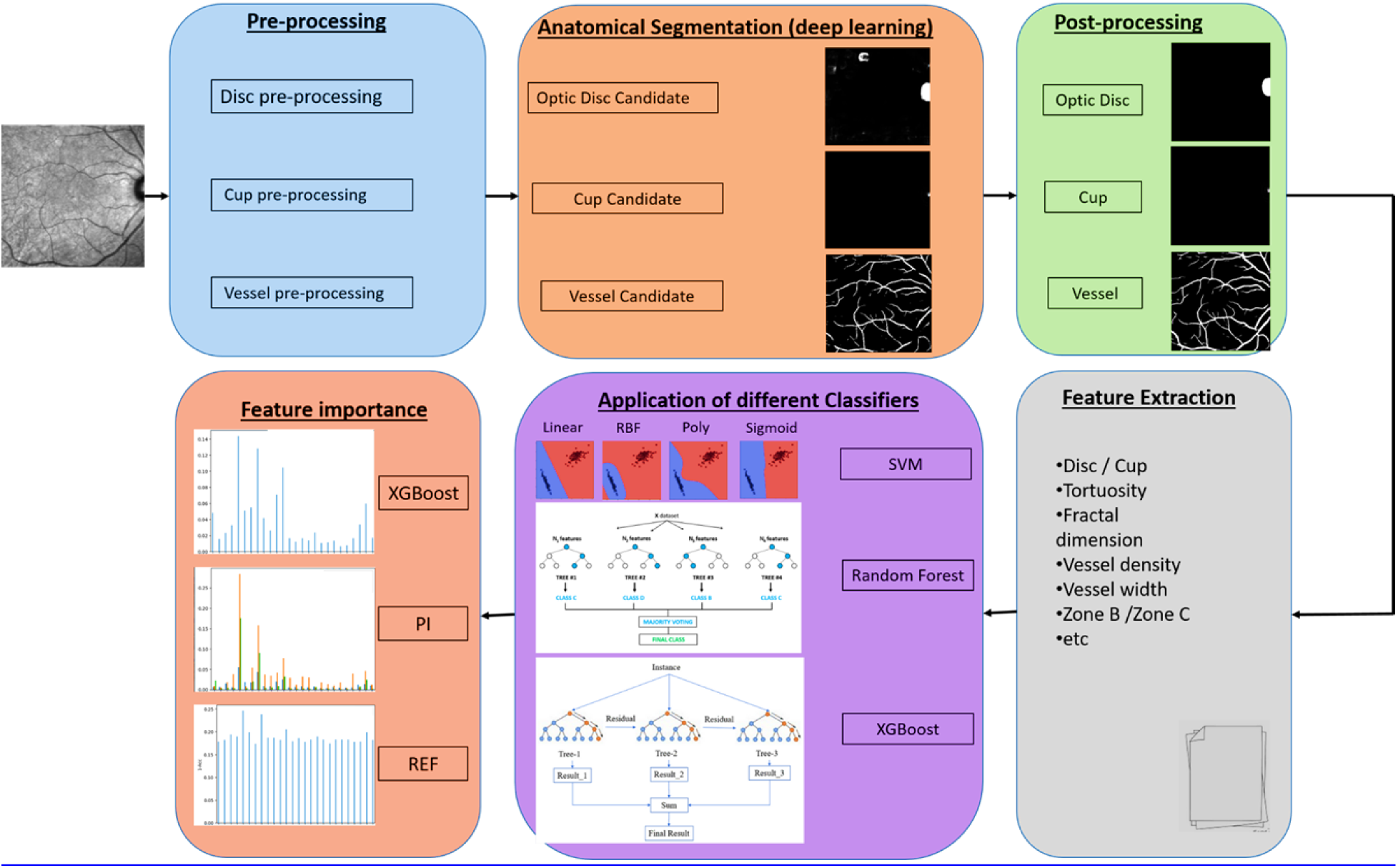
Overview of the proposed method to analyze the morphological changes in SLO images related to MS

### 2.1. Dataset

We utilized two independent datasets, namely the Isfahan and Johns Hopkins datasets, which included OCT scans and SLO images obtained from individuals with MS and HC subjects. These datasets were captured using the Heidelberg SPECTRALIS SD-OCT device (Heidelberg Engineering, Heidelberg, Germany). The Isfahan dataset, acquired from a prior study by Ashtari et al. [18], comprised a total of 282 SLO images. Among these, 146 images were from 35 patients with MS, and 136 images were from 71 HC individuals. The study took place between April 2017 and March 2019 at the Kashani Comprehensive MS Center in Isfahan, Iran, which serves as a primary referral center for MS in the region. The publicly available Johns Hopkins dataset included SLO and OCT images from the right eyes of 35 individuals, consisting of 14 HCs and 21 patients with MS [19]. Notably, this dataset exhibited demographic and clinical differences compared to those in the Isfahan dataset."

#### 2.1.1. Test, validation, and train data splitting

The two distinct datasets discussed in the previous section were combined into a unified dataset by stacking them together. To prevent any leakage among test, validation, and training samples, a subject-wise approach was adopted, wherein all images associated with a specific subject were exclusively designated for either the test, validation, or training set [20].

For the division of the test dataset, stochastic matching based on age and gender was employed initially to address potential confounding variables between the HC and MS groups. This was done before proceeding with the subsequent separation of training and validation data [21]. Initially, 20% of subjects with MS in the dataset were randomly chosen and designated as the test dataset. For each selected MS case, an HC patient with the closest age and the same gender was also included in the age-gender matching test dataset.

Following this, the splitting of train and validation data was conducted using k-fold cross-validation (CV) on the remaining patients. This approach, preferred over a random split for its completeness and generalization, ensures that the entire dataset is utilized for training. In this method, predictive models are evaluated by dividing the dataset into k folds and training and evaluating the model k times, each time using a different fold as the validation set. Moreover, to maintain an equal proportion of certain labels (MS or HC) in each fold, stratified sampling was employed.

#### 2.1.2. Data augmentation

Data augmentation is a well-known technique in ML studies used to artificially expand the size of a limited training dataset to mitigate the risk of over-fitting. This involves making minor alterations to the existing training dataset to create new and plausible examples. In this study, several geometric and color space transformations were performed, including vertical and horizontal flips, height and width shifts within the range of ±5 pixels, rotation within the range of ±15 degree, and adjustments to image brightness in the range of 0.8 to 1.5.

### 2.2. Feature Extraction

To identify alternative and tangible clinical characteristics in MS SLO images, we employed basic machine learning models. These models were trained using manually extracted features related to the optic disc, cup, and vascular morphology, rather than utilizing deep learning models. This approach provided a clearer insight into the clinical significance of different features in the classification of MS and HC. To accomplish this, the initial steps involved the segmentation of the optic disc, cup, and vessels from the SLO images, followed by the extraction of clinically relevant features.

#### 2.2.1. Anatomical Segmentation

As the quantity of SLO images at our disposal was limited, and we lacked corresponding ground truth for these images, we leveraged the pre-trained anatomical segmentation models outlined in [22]. These models had been trained on public datasets comprising retinal fundus photographs with associated ground truth. However, the inherent differences in imaging modalities between SLO images (characterized as monochromatic fundus imaging with single-wavelength laser light) and fundus photographs (using red, green, and blue wavebands for imaging light) [23] may lead to errors in the initially proposed deep learning models in [22] when our dataset is applied as test data for these models. Consequently, we implemented several pre-processing and post-processing techniques to tailor the segmentation for the optic disc, cup, and vessels in SLO images. Following sections provide a detailed explanation of the models utilized for the segmentation of optic disc, cup, and vessels, respectively.

##### 2.2.1.1. Optic Disc Segmentation

To achieve optic disc localization and segmentation, we employed the pre-trained LW-Net model outlined in [22]. However, as mentioned earlier, adjustments were made to the model outputs to make them applicable to monochromatic SLO images. The optic disc segmentation phase utilized in this study comprises three primary sub-stages: pre-processing, identification of optic disc candidates, and post-processing. Detailed explanations of each stage are provided below.

###### Pre-Processing

Typically, the optic disc presents as a bright yellowish or white area in color fundus images (brighter than the background when the image is converted to grayscale), but it appears as a dark region in SLO images (darker than the background). To address this difference, in the initial stage, the pixel intensity values in SLO images were inverted, swapping black pixels with white and vice versa. As a result of this adjustment, the optic disc manifested as bright regions in the SLO images.

In this study, many SLO images exhibited variations in brightness and uneven illuminations, potentially leading to inaccuracies in identifying optic disc candidates in subsequent stages. To address this issue, we employed a specific SLO image as a reference, characterized by a clearly discernible optic disc and minimal intensity fluctuations Figure 2 shows the SLO image selected as a reference. As can be seen, this is an SLO image where the optic disc is clearly visible in comparison to the background and the variations in background intensity are also negligible. Subsequently, in order to manipulate the intensity distributions and mitigate contrast level variations in other SLO images, their histograms were matched with the histogram of the chosen reference image.

**Figure 2.**
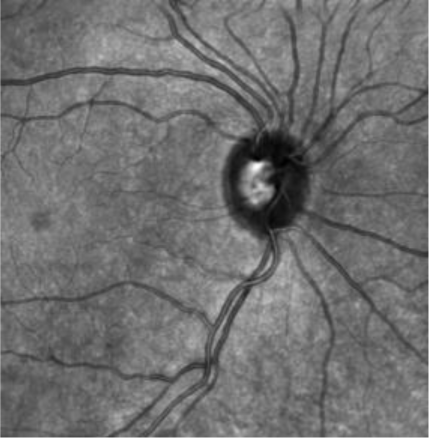
The SLO image selected as the reference image for the histogram matching algorithm.

In SLO images, the central reflection of blood vessels is observable on both arteries and veins [23]. This reflection tends to be more pronounced on arteries than on veins, especially when working with color fundus images. These central reflections on veins, which are the thicker, clear vessels around the optic disc, were observed as dark strips with an intensity level nearly similar to that of the cup in some of the inversed SLO images derived from the previous step. This phenomenon poses the risk of generating inaccurate optic disc candidates in subsequent stages. To mitigate this effect, two morphological closing and opening operation with a rectangular structure element were applied to the intensity of the inversed SLO images obtained from the previous stage. Figure 3 represents these central reflections as dark strips on veins, along with the image resulting from the removal of their effects —depicted in the left and right images, respectively.

**Figure 3.**
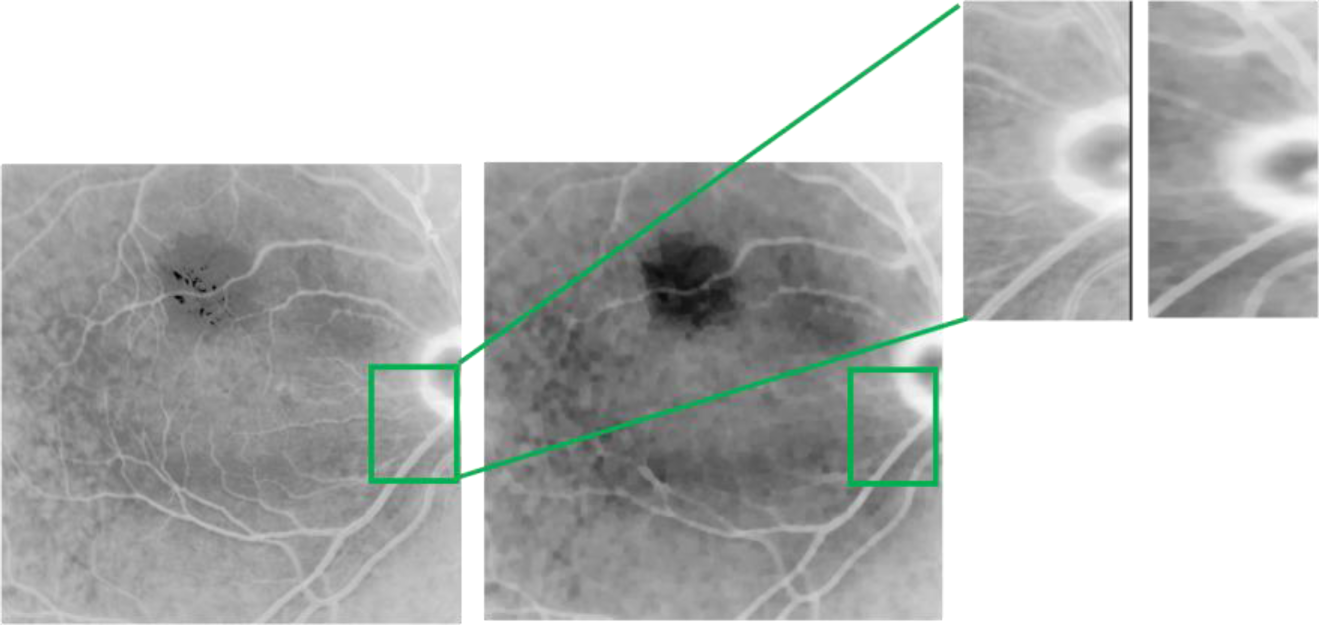
Inversed matched SLO images with central reflection as dark strip on veins, the left image, and the image resulted from applying morphological closing and opening operation, the right image. The windows containing veins around the optic disc are also magnified.

Finally, every SLO image was resized to (512,512) to accommodate the large batch size required by the LW-Net model [22]. The application of the LW-Net model in calculating optic disc candidates is explained in detail in the following section.

###### Optic Disc Candidates

To identify candidate regions for the optic disc in the SLO images after undergoing pre-processing, we utilized the LW-Net model, which comprises two U-Nets as described in [22]. However, as mentioned earlier, the monochromatic nature of SLO images, in contrast to the 3 color channels of fundus images used in [22] as a dataset, necessitated adjustments and modifications in the output of the second U-Net within LW-Net.

Initially, the SLO images were utilized as inputs for the encoder, which corresponds to the first U-Net in the LW-Net. The outputs of the decoder, associated with the second U-Net in the LW-Net, underwent classification into three categories: background, optic disc and cup. We specifically regarded the second channel resulting from the decoder as the segmentation map for disc candidates. Finally, we examined these candidates to delineate the optic disc area.

###### Post Processing

During this stage, the binary images containing optic disc candidates underwent evaluation based on distinct characteristics such as shape, bounding box, and coordinates. Hence, the optic disc was segmented utilizing the following five main sub-stages:

- In order to eliminate noise pixels between two or more candidates and to isolate each candidate, particularly in low-quality SLO images exhibiting intensity variations despite the application of the histogram matching method, morphological closing and opening operations, using a structure element in the shape of ellipse, were conducted on the binary images that encompass the candidates.
- Since the optic disc appears as bright areas in inverted images (generated during the pre-processing phase), dark regions cannot be identified as the optic disc. To filter out candidate areas with a low probability of being the optic disc, candidates with a mean intensity lower than a specific threshold were excluded. This threshold was determined based on the mean intensity values of all candidates in each image.
- Candidate regions in each image with an area smaller than a certain threshold were excluded (2300 pixels for candidates located on both sides of the images and 3000 pixels for those located near the center of the images).
- The shape and area of the remained candidates were determined using connected component analysis, and those that with a line shape or a low width-to-length ratio in their bounding box were eliminated. For candidate regions located near the center of the images, those with a low length-to-width ratio in their bounding box were also removed. Ultimately, the candidate with maximum area was designated as the final optic disc candidate.
- The final optic disc candidate underwent a blob detection algorithm to delineate the boundary of the optic disc. While the optic disc typically appears nearly circular, an ellipse transform was employed because in some images only one arc of the optic disc may be visible. The algorithm used to calculate the boundary and radius of the Optic disc candidate positioned on the sides of the SLO images is summarized in supplementary material on page 1.

##### 2.2.1.2. Cup Segmentation

To localize and segment the cup in the SLO images, we utilized the outcome obtained from optic disc segmentation stage. Initially, a window surrounding the segmented optic disc in the original data (not the inversed one) was considered and then provided as input to the first U-Net in the LW-Net model described in [22]. The first channel of the output of the LW-Net model was treated as a segmentation map for cup candidates.

Subsequently, the mean intensity value of each candidate was compared to a certain threshold (the mean intensity of all candidates in each image), and if the intensity value of the candidate was smaller than the threshold, it was excluded. Then, within each image, the center of bounding box of every remaining candidate and its distance from the center of segmented optic disc were computed. The cup candidate with the smallest distance was chosen as the final candidate. However, it must adhere to three main criteria (unless it should be omitted):

i. The bounding box of the candidate should be entirely situated within the optic disc boundary.
ii. The candidate’s width-to-length or length-to-width ratio must be less than 2 (as the cup does not have a narrow oval shape.).
iii. The area of the candidate must have been greater than a certain threshold (700 in this work).

Ultimately, should a cup candidate be present, the boundary of the cup would be established using an ellipse transform, akin to the method outlined in the optic disc segmentation section.

##### 2.2.1.3. Vessel Segmentation

To segment the vessels in SLO images, we employed the pre-trained method proposed in [22], complemented by a post-processing step that played a crucial and effective role in accurately segmenting blood vessels. These two main stages are explained in detail in bellow.

###### Binary Vessel Segmentation Map

Because of uneven illumination and intensity variations among the SLO images in the utilized dataset, our initial step involved employing the histogram matching algorithm using a proper reference image. This reference image, displayed in the supplementary material on page 2, is characterized by minimal intensity changes, served to eliminate variations in brightness and prevent the segmentation of false candidate pixels that may represent vessels.

Afterward, we employed the SEGAN model designed in [22], which is a variant of U-Net comprising a segmentor and a discriminator trained using an adversarial learning strategy. The SLO images, having undergone histogram matching, were initially resized to (912,912) to alleviate the computational demands before being used as inputs for the SEGAN model. The output from the discriminator provided a segmentation map, where each pixel represented the likelihood of being a blood vessel. In this study, pixels with a likelihood greater than 0.3 were chosen to create the binary representation of blood vessels.

###### Post Processing

Since we used the proposed SEGAN model [22] trained by six public dataset containing fundus images and only tested the model on our dataset, there were some discontinuities in the segmented blood vessels resulting from the proposed SEGAN model. To solve this problem, we used a post-processing step and took advantage of two useful algorithms, a region growing method and a missing algorithm [24], to fill discontinuous parts of the segmented blood vessels.

At the first stage, to overcome the discontinuities occurring in the segmented vessels, especially along the big blood vessels, we utilized the region growing algorithm which starts with some seed pixels in an image and grows regions from them by iteratively adding unassigned neighboring pixels that satisfy some certain criterion with the existing regions of the seed pixels found in. Figure 4 represents a region growing algorithm. For this, we first computed an image skeleton of the blood vessels segmented from previous step and then construct an undirected vessel graph and considered terminal or end nodes, pixels that belong to vessel skeleton and have only one neighboring skeleton pixel.

**Figure 4.**
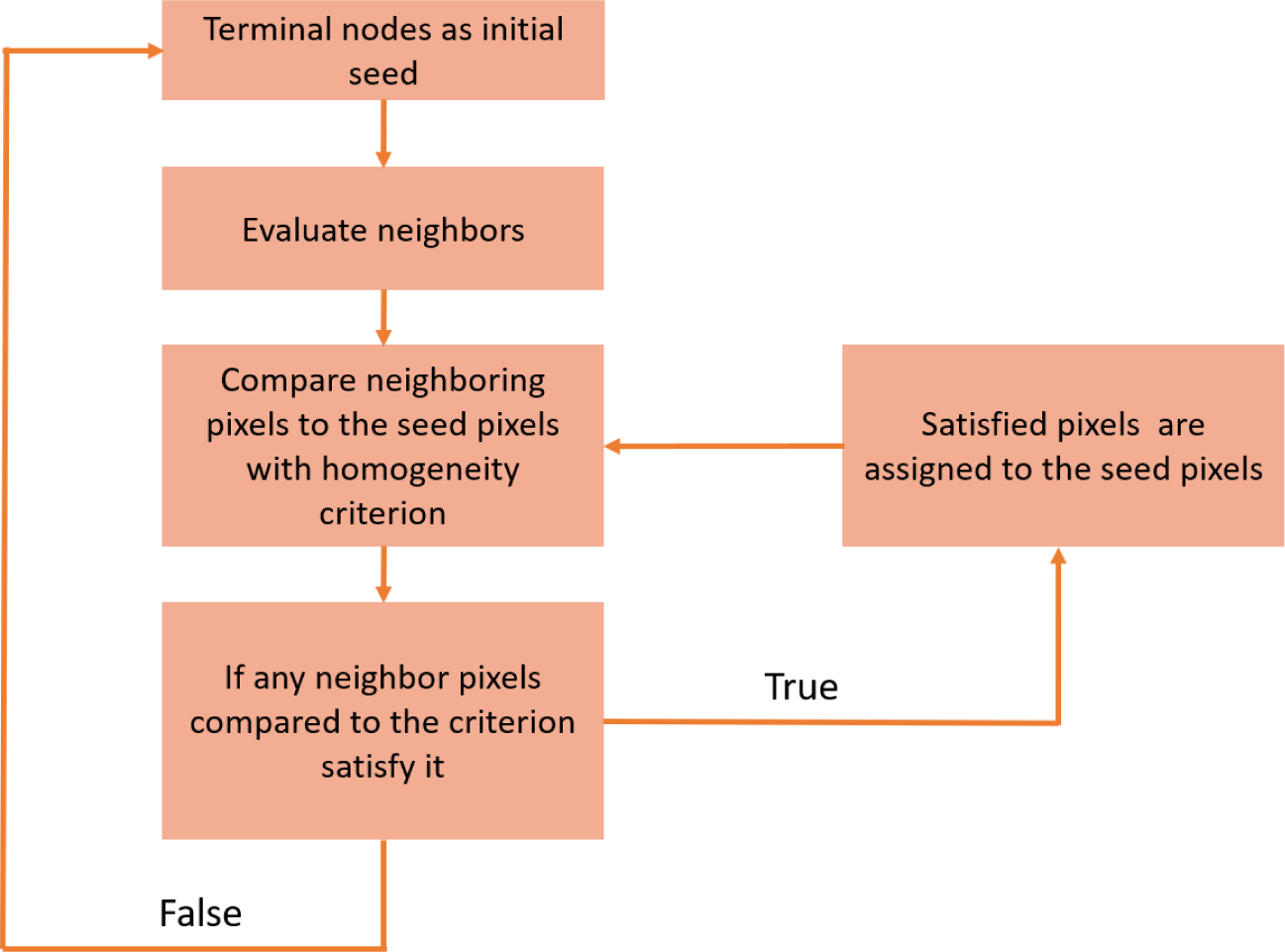
The region growing algorithm used for vessel segmentation.

To implement the region growing algorithm, for each terminal point that its intensity is greater than a certain value, 40 for this work, the intensity of its neighboring pixels not belonging to segmented vessels were considered and those whose brightness difference with a certain threshold were less than a predetermined value,10 for this work, were selected as candidate points and the candidate point with lowest brightness difference was selected as new seed. The threshold value for selecting candidate points is the intensity of the terminal point and will be updated with mean intensity of the terminal point and the new selected seed. For each selected new seed, the above algorithm was repeated until no neighboring point satisfied the criterion. Algorithm 1 states our proposed region growing approach in detail in which p_i_ represent ith terminal point, S_1_ is a set of neighboring points satisfying a certain criterion that was explained above and finally P_S1,i_ represent ith point in S1.

###### Algorithm 1. The region growing model proposed for addressing discontinuities in binary vessel map

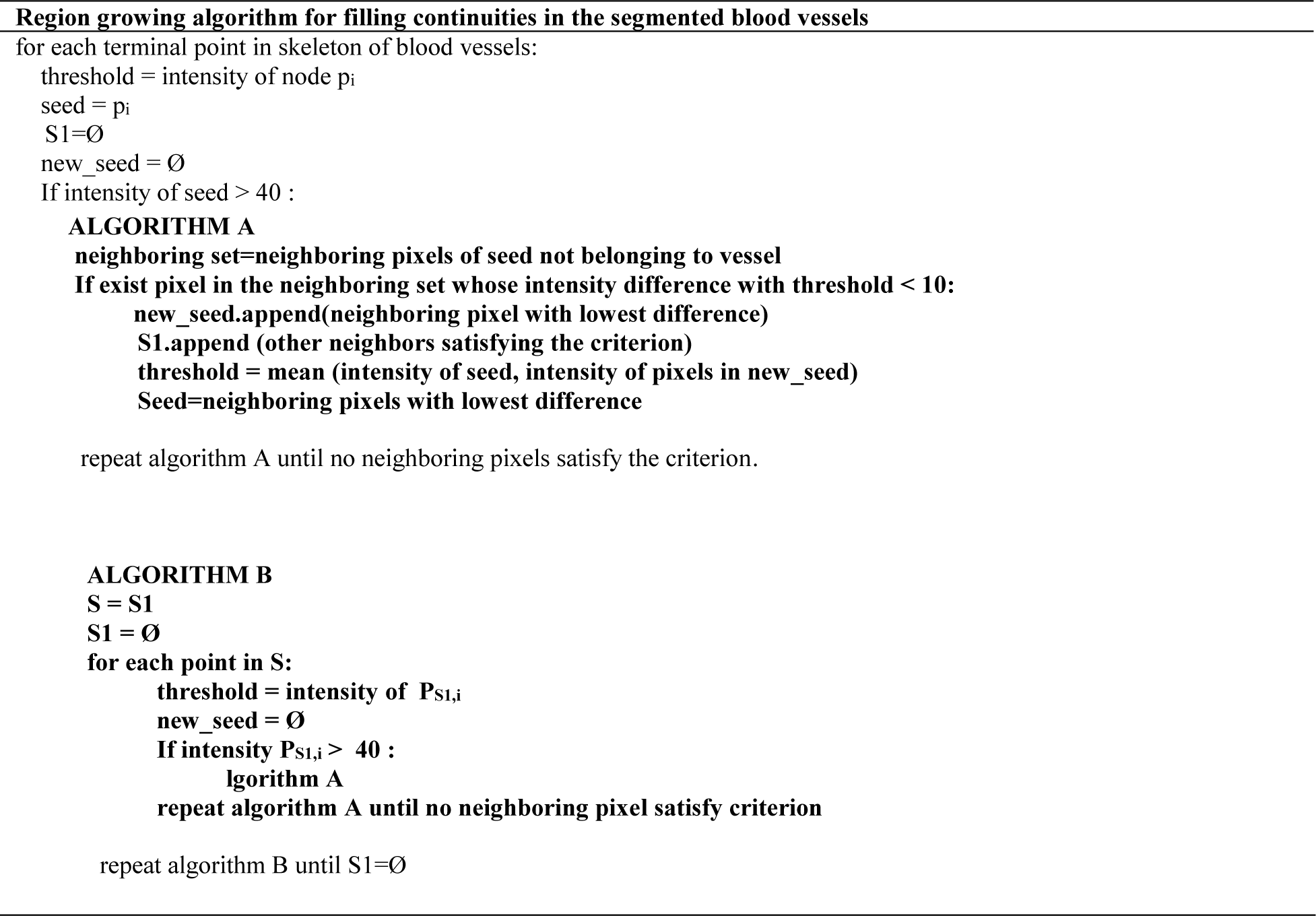

In the next step, we used the missing algorithm proposed [24] to compensate for some small discontinuities left over from the first step. The proposed missing algorithm corrected the small disconnected segments in vessel segmentation by taking advantage of the vessel graph and determining the landmark points on it.

#### 2.2.2. Feature Measurement

In this work, a series of clinically relevant manual features from segmented optic disc, cup and blood vessels [22] was calculated to classify individuals in 2 classes, i.e. MS and HC, as summarized in table 1,

**Table 1.** The feature set extracted from Optic Disc, Cup and blood vessels in the SLO images.

| **Manual features extracted from SLO images** |  |
| --- | --- |
| Features from Optic Disc / Cup | Disc radius<br>Cup radius<br>Disc radius-to-Cup radius<br>Disc location |
| Vascular features from whole image | Average width<br>Fractal dimension<br>Vessel density<br>Vessel intensity<br>Distance measure tortousity<br>Squared curvature tortousity<br>Tortousity density |
| Vascular features from Zone B and Zone C | Average width<br>Fractal dimension<br>Vessel density<br>Linear regression tortousity<br>Distance measure tortousity<br>Squared curvature tortousity<br>Tortousity density |

Radius of disc, cup and ratio between them, plus location of disc in the SLO images were the feature set measured form Optic Disc and Cup in the training, validation and test datasets.

Also, average width, a measure describing average change of vessels width, fractal dimension value [25], a metric of vessel complexity, vessel density indicating the ratio between the area of vessels to the whole image, vessel intensity, the ratio between intensity of vessels to intensity of whole image, plus three different methods for calculating vessel tortuosity, including distance measure tortousity [26], providing a ratio of the actual path length to the linear distance between curve endpoints, Squared curvature tortuosity [26] and tortuosity density [27], were computed as vascular features from the whole image.

Furthermore, Vascular features in standard regions of SLO images including zone B, the region between 2 and 3 times the radius of optic disc from the center of optic disc, and zone C, the area between 2 and 5 times the radius of optic disc from the center of optic disc, were considered as other manual features [28]. A figure representing zone B and zone C from the center of optic disc is shown in the Supplementary Material on page 3. These features consist of average width, fractal dimension value, vessel density, linear regression tortuosity, a coefficient measuring the linearity of each segment of vessel, distance measure tortousity, Squared curvature tortuosity and tortuosity density.

### 2.3. Classification

As mentioned before, the aim of this work was to determine important, valuable and useful features that clinically differentiate and discriminate MS SLO images from HC ones. For this purpose, we employed classification application.

First, to evaluate the features extracted from the Optic Disc, Cup and blood vessels of the SLO images, we used simple machine learning models including, SVM, Random Forest (RF) and XGBoost classifiers, and then measured the importance of each feature to determine which features are discriminative and tangible to classify individuals in two classes, namely MS and HC.

Each of the aforementioned classifiers was trained by concatenating the extracted features from training data of each fold, constructed by k-fold cross validation, and their augmented SLO images, and then tested on the features extracted from the validation data in those folds. After that, a set of metrics were employed to evaluate the performance of each classifier and their average on k validation folds was reported as the classifiers performance (k=5 in this work). Then, the feature importance was calculated as a score representing the importance of each feature in the set extracted features, providing a clear insight into how effective is each feature in the behavior of the predictive machine learning model.

#### 2.3.1. SVM Classifier

At the first step, we utilized the SVM classifiers, a kind of supervised machine learning method for classification, regression and outlier detection tasks, with different kernel functions, including linear, radial basis function (RBF), polynomial (poly) and sigmoid function to classify the features extracted from SLO images in two classes, MS and HC.

Furthermore, to search for optimal SVM hyperparameters, including regularization parameter (C) for linear, RBF, poly and sigmoid functions, kernel coefficient (gamma) for RBF, sigmoid and poly kernels and degree of polynomial kernel function, Optuna, an automatic hyperparameter optimization software framework particularly designed for machine learning was utilized [29].

#### 2.3.2. Random Forest

A Random Forest (RF) or Random decision forest classifier, a supervised learning algorithm based on bagging technique consisting of a number of decision tree classifiers on various sub-samples of the dataset that uses averaging to improve the predictive accuracy and control over-fitting, was another simple classifier that was used in this work to evaluate the extracted features from the SLO images to classify them into two classes, MS and HC. Hyperparameters for RF classifier, including the number of trees in the forest, the maximum depth of the tree, the minimum number of samples required to split an internal node and the minimum number of samples required to be at a leaf node, were also determined and optimized by Optuna optimization framework [30].

#### 2.3.3. XGBoost classifier

Also, we used XGBoost classifier, an implementation of boosting technique that sequentially creates decision trees and each tree improves upon the mistakes of the previous one, to classify MS and HC SLO images. XGBoost is an ensemble learning method that offers a systematic solution to combine the predictive power of multiple learners. A model whose parameters adjust itself (XGBoost) will learn better than one with a fixed set of parameters for the entire ensemble (Random Forest). Then, we took advantage of Optuna optimization software to get optimal hyperparameters for XGBoost classifier [30].

#### 2.3.4. Evaluation of Classifiers

To evaluate the performance of aforementioned classifiers, a set of metrics including Accuracy (ACC), Sensitivity (SE), Specificity (SP), Precision (PR) and F1-score (harmonic mean between precision and recall) was employed with the following mathematical formula:

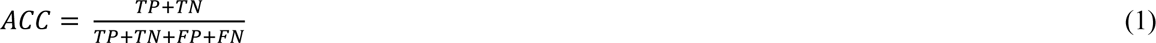

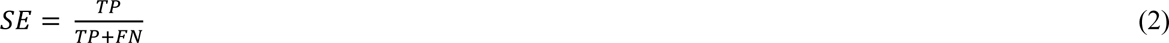

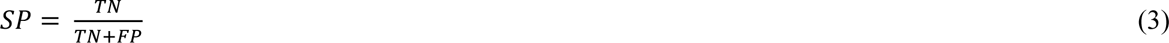

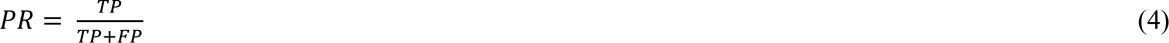

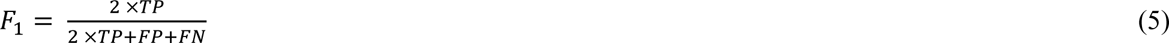

Where TP, FN, TN, and FP are the metrics representing true positive, false negative, true negative and False positives, respectively. Furthermore, receiver operating characteristics (ROC), showing the relationship between true positives and false positives, and precision recall (PR), illustrating the trade-off between precision and recall, were utilized to measure the performance of the above classifiers.

#### 2.3.5. Feature Importance

In general, feature importance provides a score indicating how useful and important each feature in a feature set contributes to the model prediction and therefore gives a highly compressed and global insight into the behavior of the model. The importance calculated for each feature in a feature set also allows features to be ranked and compared with each other. In this study, we used three different methods (XGBoost classifier, recursive feature elimination, RFE, and permutation importance, PI) for measuring importance of each feature in the feature set obtained from section 2.2.2.

One advantage of using gradient boosting is that it can automatically provide estimates of feature importance from a trained predictive model and also is relatively simple to measure the importance scores for each feature after the boosted trees are constructed. The feature importance for a single decision tree is calculated by the amount by which each split point improves the performance measure, weighted by the number of observations that the node is responsible for, and then the importance of that feature is averaged across all decision tress in the model [31].

Recursive feature elimination (RFE)[32], a wrapper-type feature selection method, was another method that we used for feature selection. RFE is a popular feature selection method that is easy to use and effective in selecting those features in the training data set that are more or less relevant in predicting the target variable. REF works by searching a subset of features starting with all features in the training set and then eliminating features one by one until the desired number remained. Finally, the important features obtained from the above two methods for feature selection were compared.

Permutation importance (PI) [33] was the third algorithm used in this study for calculating feature importance. The concept behind PI is really straightforward: it calculates the importance of a single feature by increasing the model error. In this manner, a feature is important if shuffling its value increase model error, meaning that the feature plays an important role for the model. In the same way, a feature is unimportant if shuffling its value leaves the model error unchanged.

## 3. Results

All the experiments in this study were performed by using Python programming language in backend Torch platform in python 3.7 software environment.

Overall, 314 SLO images from stacking two independent datasets, Isfahan and Johns Hopkins datasets, consisting of 85 and 53 subject groups for HC and MS individuals, respectively, were used. To construct test dataset based on age-gender matching algorithm explained in section 2.1.1, 10 MS patients and HC individuals with the nearest age were selected. The remaining images from our dataset were utilized for splitting data into train and validation sets using k-fold CV (K=5). All images were resized to 512 × 512 × 1with intensity level values in range [0, 256). Furthermore, all SLO images which belong to the left eyes were flipped.

### 3.1 Segmentation

We took advantage of the proposed LW-Net model [22], trained by REFUGE dataset containing retinal fundus photographs, to segment Optic Disc, Cup and vessels in SLO images. But, as mentioned in section 2.2.1, to obtain an accurate segmentation from the SLO images in our dataset, we modified the proposed method and used pre-processing and post-processing stages as two effective steps.

Visualization results for optic disc and cup segmentations are represented in figure 5. In this study, we were able to segment the optic disc even when it was located on the left or right side of the image and only an arc of its circular border was visible.

**Figure 5.**
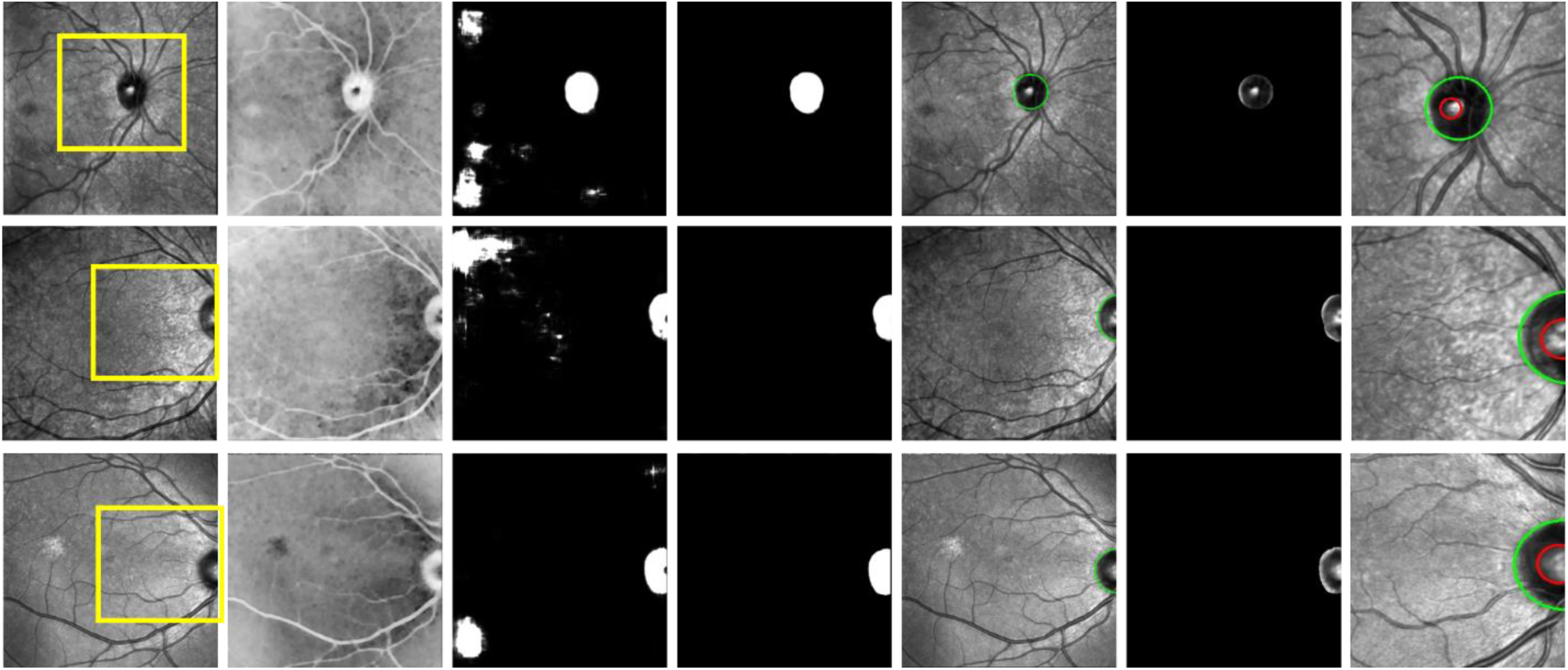
visualization results for Optic Disc and cup segmentations, including pre-processing step for disc segmentation (the second column), optic disc candidates (the third column), post-processing step for disc segmentation (the fourth and fifth columns), windows surrounding the segmented optic disc (the sixth column) and finally the optic disc and cup segmented by green and red circles, respectively (the last column). The last column shows the zoomed state of the yellow window on the SLO images in the first column.

The segmentation results for blood vessels in the SLO images are also shown in figure 6. The results for region growing and missing algorithms are displayed in the third and fifth columns, respectively. The fourth and sixth columns represent the results of these two stages for a small window of the images.

**Figure 6.**
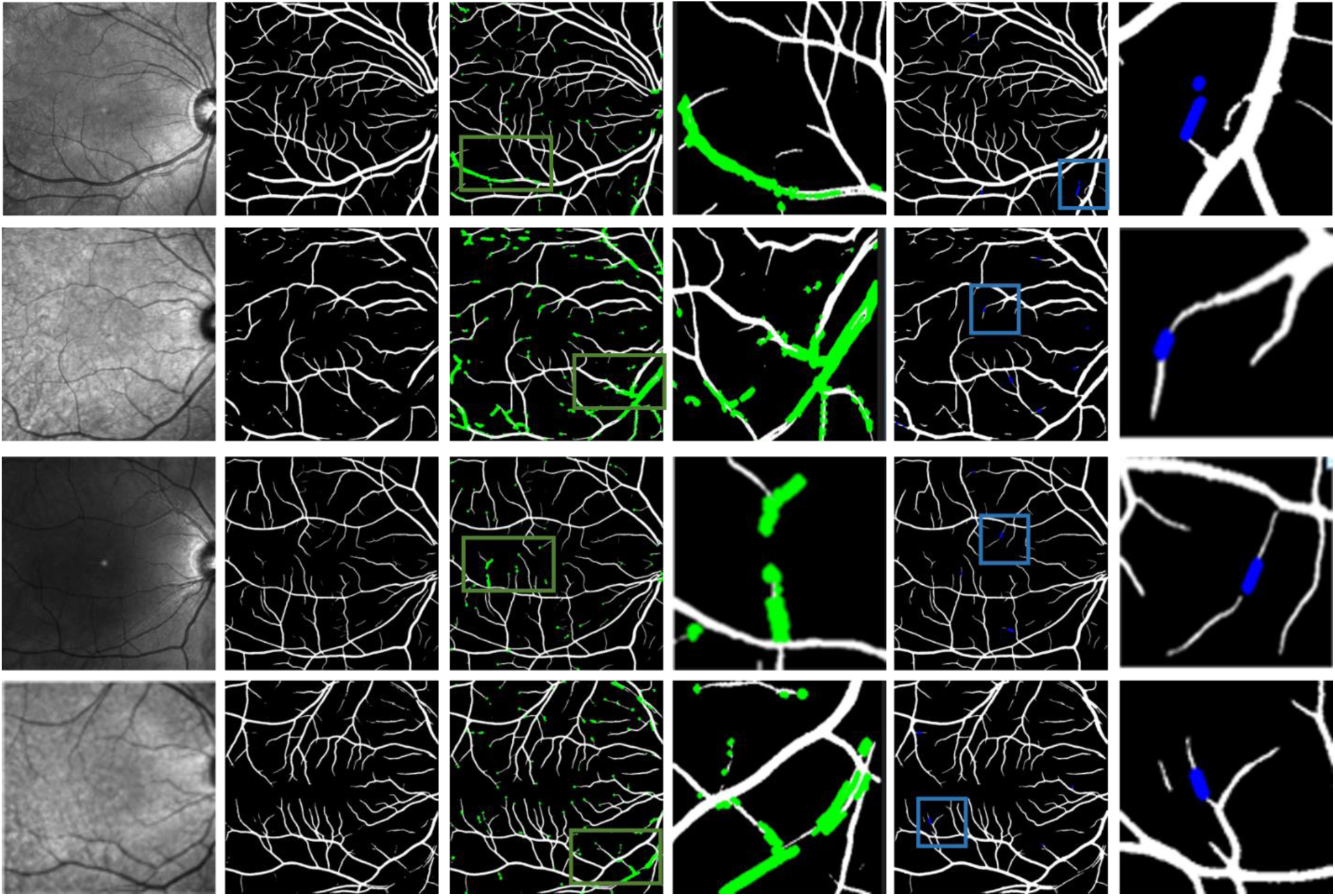
visualization results for vessel segmentation, including the proposed algorithm [22] (the second column), the region growing and missing algorithms (the third and fifth columns)

### 3.2. Classification

To evaluate the extracted manual features from the SLO images, a set of simple machine learning classifiers consisting of SVM, RF and XGBoost algorithms were utilized for classifying MS and HC. The XGBoost classifier showed the best performance with a mean ACC of 83% (SP=0.85, SE=0.79, F1=0.83, AUROC=0.84, AUPRC=0.87) as the winner classifier. Table 2 and 3 represent the performance of the used classifiers, and their corresponding hyperparameters optimized by Optuna, respectively.

**Table 2.** Performance metrics of three simple machine learning models (SVM, RF and XGBoost classifiers) for classification of MS using SLO images. Best results are bolded, revealing that XGBoost classifier is the wining classifier.

| Model | ACC | SP | SE | F1 | AUROC | AUPRC |
| --- | --- | --- | --- | --- | --- | --- |
| **SVM**<br>**(kernel : RBF)** | 0.80 | 0.90 | 0.66 | 0.79 | 0.84 | 0.85 |
| **RF** | 0.82 | 0.84 | 0.77 | 0.81 | 0.83 | 0.85 |
| **XGBoost** | **0.83** | **0.85** | **0.79** | **0.83** | **0.84** | **0.87** |

**Table 3.** The tuned optimal hyper parameters by Optuna for SVM, RF and XGBoost classifiers used for classification MS using SLO images.

| **Model** | **Optimal hyper parameters** |
| --- | --- |
| SVM (kernel : RBF) | C =91.42<br>Gamma = 0.008 |
| RF | max_depth = 16<br>n_estimators = 137<br>min_samples_split = 7<br>min_samples_leaf = 3<br>criterion = gini |
| XGBoost | objective = binary :logistic<br>tree method = auto<br>booster = dart<br>lambda = 1e-7<br>alpha = 0.5<br>subsample = 0.99<br>colsample_bytree = 0.79<br>max_depth = 4<br>min_child_weight = 3<br>eta = 0.34<br>gamma = 0.2<br>grow_policy = deptwise<br>sample_type = forest<br>rate_drop = 0.96<br>skip_drop = 0.3 |

Figure 7 also shows the confusion matrix for all the three classifiers to visualize and summarize the performance of the used machine learning classifiers.

**Figure 7.**
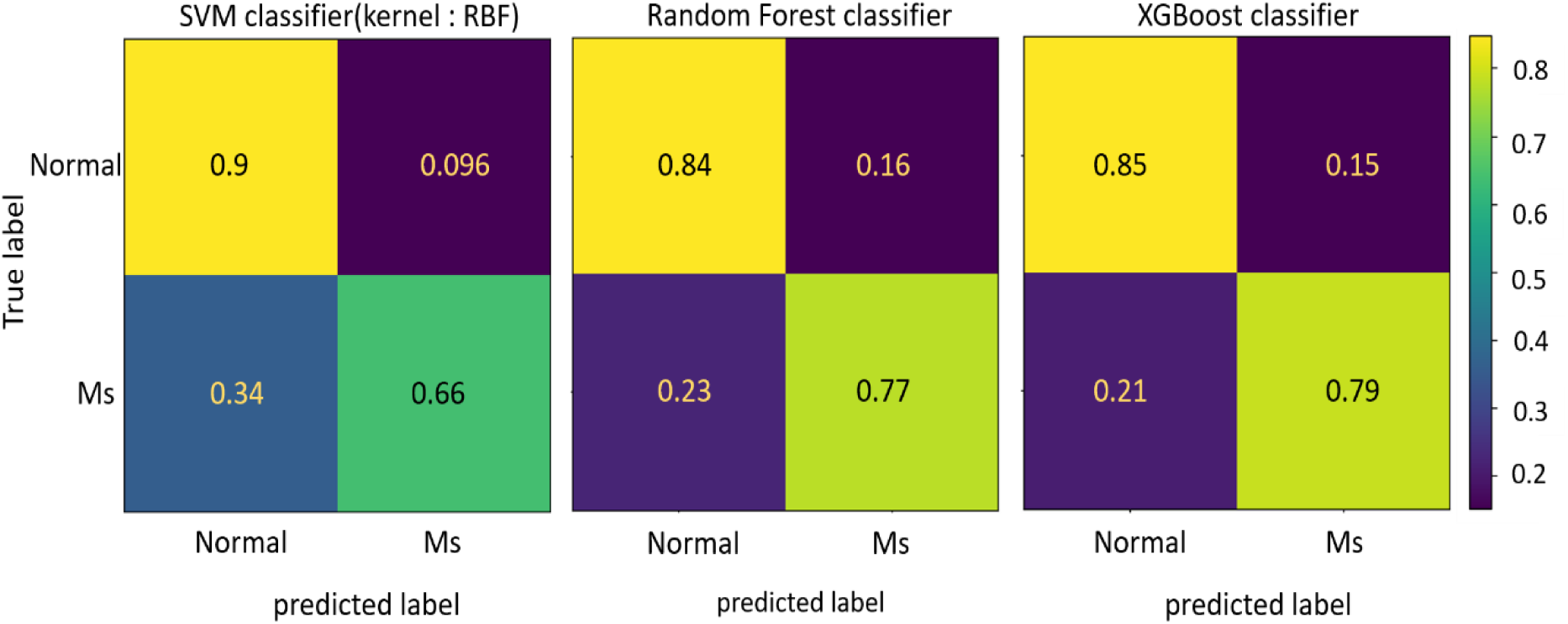
confusion matrix for three machine learning models including SVM, RF, XGBoost classifiers.

Then, we tested the trained XGBoost classifier on age-gender matching test dataset. The designed XGBoost classifier resulted in a mean ACC of 0.72 (SP=0.86, SE=0.65, F1=0.73, AUROC=0.83, AUPRC=0.92).

### 3.3. Feature Evaluation

We measured feature importance for the build feature set in section 2.2.2 to get a better understanding of what features were more discriminative and useful for HC and MS classification. Since XGBoost was the classifier with the highest performance, we first took this advantage and used a built-in attribute of XGBoost classifier to estimate the importance of each feature in the feature set obtained from the section 2.2.2. The mean importance of each feature was shown in figure 8. As can be seen, the set of features including the location of disc horizontally, fractal dimension computed from the whole image, intensity of whole vessels, tortuosity density, vessel density in zone B and vessel density from the whole image were the most important and valuable features in the construction of boosted decision trees within XGBoost classifier.

**Figure 8.**
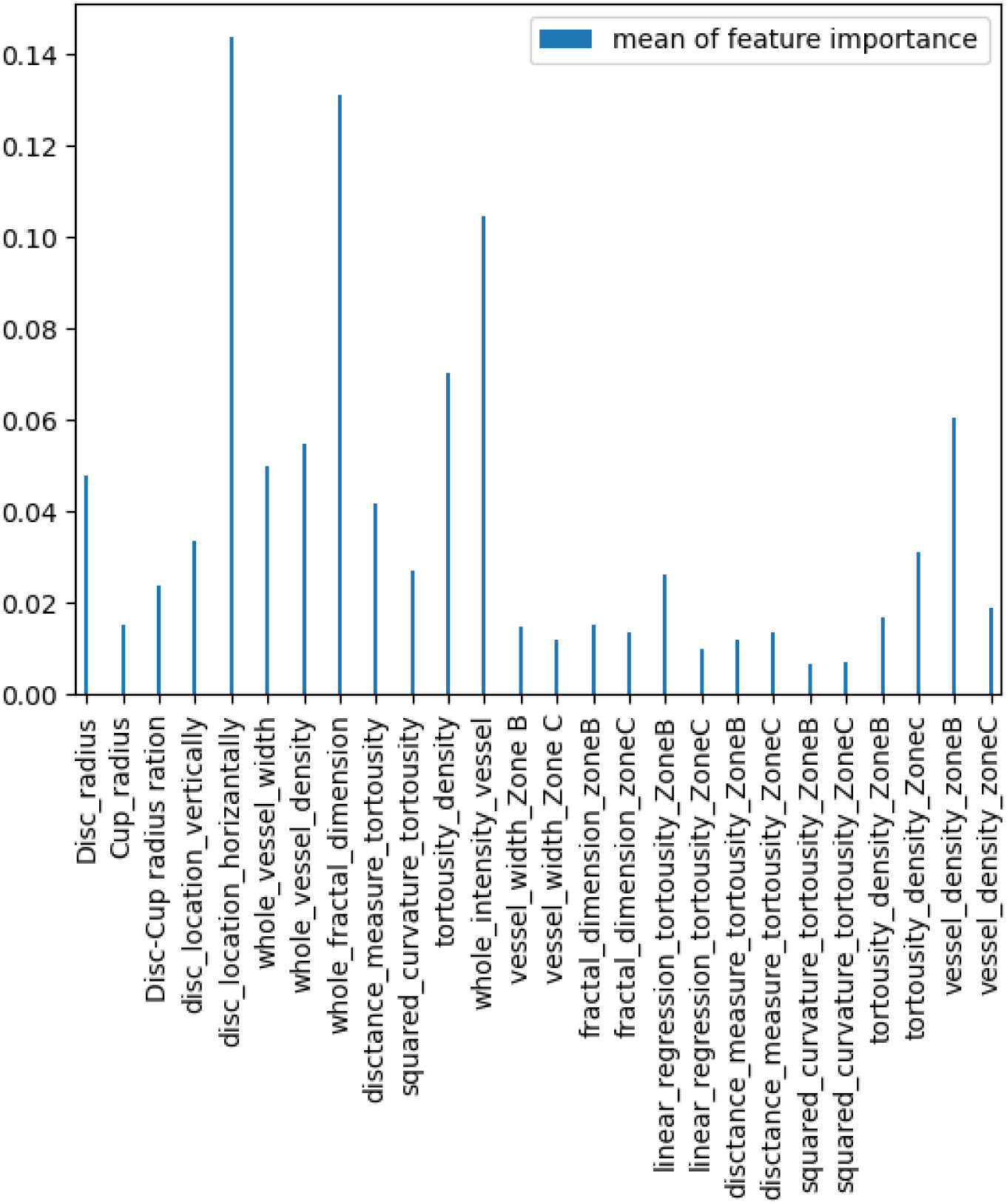
The mean of importance of each feature, on 5 folds of training data set, by using a built-in attribute of XGBoost classifier.

Figure 9 represents the distribution and the relationship between of the first six important features calculated by XGBoost classifier, giving us a better comprehension of the features and the target variable, which allows us to visualize how the most important features relate to each other. In this Figure, the X-axis and y-axis represent the first eight effective features sorted by their importance for HC and MS, respectively, and each row and column summarize the relationship between two paired features of the first eight important features.

**Figure 9.**
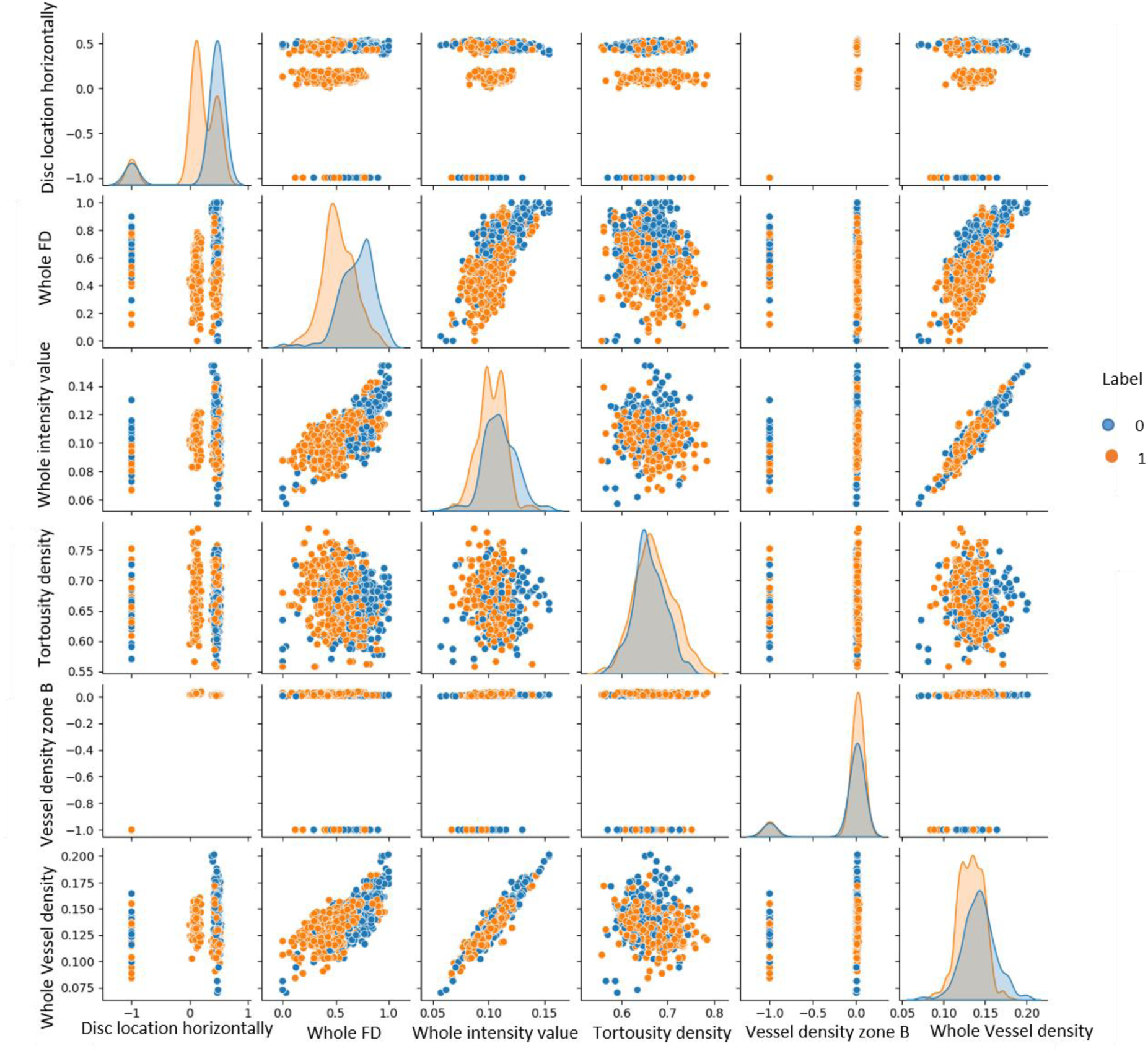
The distribution and relationship between the first sixth important features obtained by XGBoost classifier.

The importance of each feature was also measured by permutation importance (PI), as the second method) to explore the most effective features for HC and MS classification. We implemented PI method for 3 machine learning models including: Random Forest (RF), Decision Tree (DT) and XGBoost classifiers. Figure 10 displays the importance scores calculated by PI method and its mean for these machine learning models.

**Figure 10.**
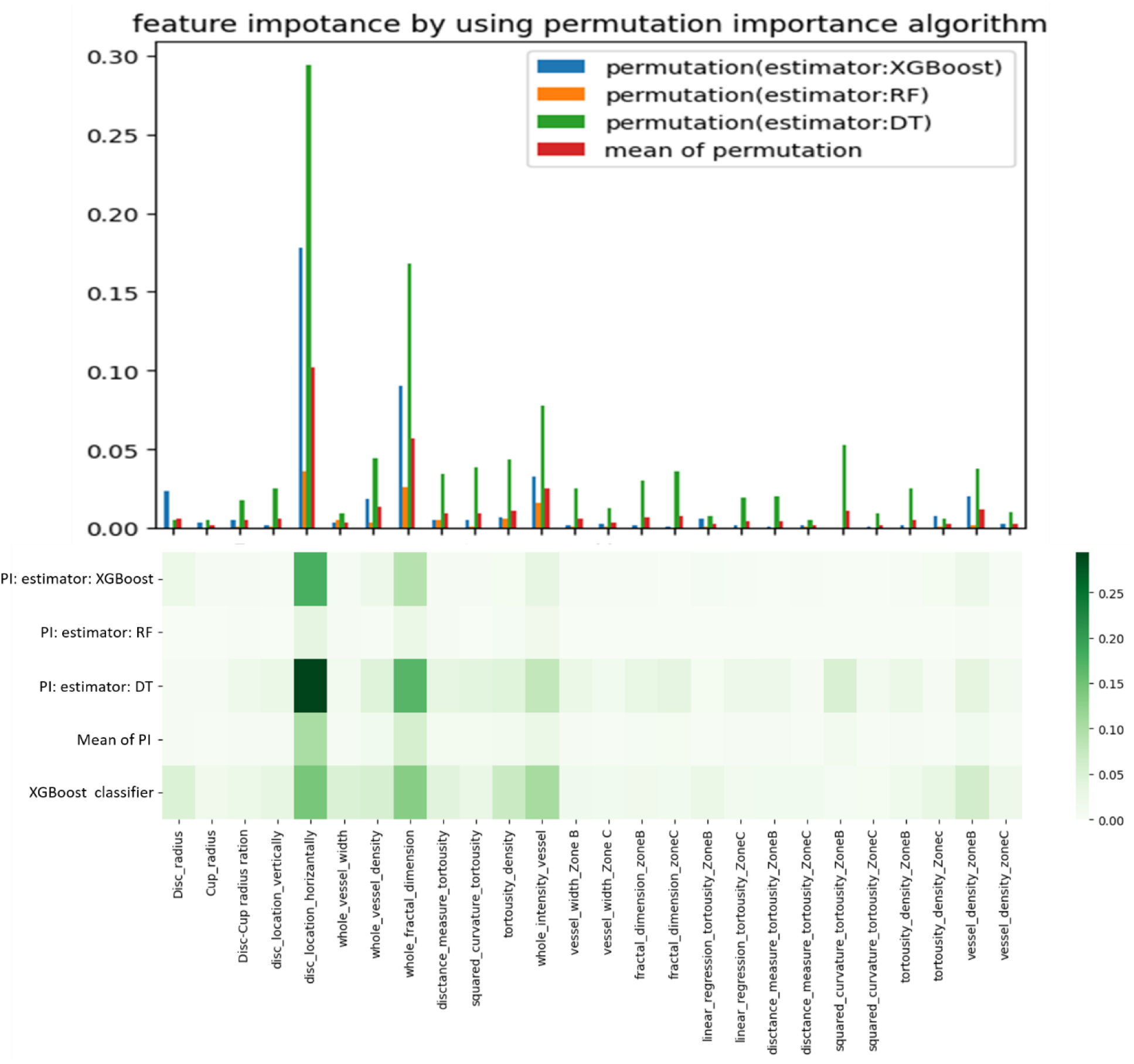
The representation of the importance calculated for each feature by PI (with 3 different classifiers), mean of PI and XGBoost classifier.

Finally, we used RFE technique as the third method to estimate the importance of each feature. As mentioned earlier, the goal of this algorithm is to recursively remove features and rank them based on the performance of a given estimator. We applied the three classifiers used in PI technique, DT, RF and XGBoost, as the estimators trained on the feature set. Figure 11 shows the feature ranking with RFE technique for the calculated feature set. As can be seen, the features named as disc location horizontally, whole fractal dimension and whole intensity value, were ranked as the three first important feature for all the three classifiers used as the estimators.

**Figure 11.**
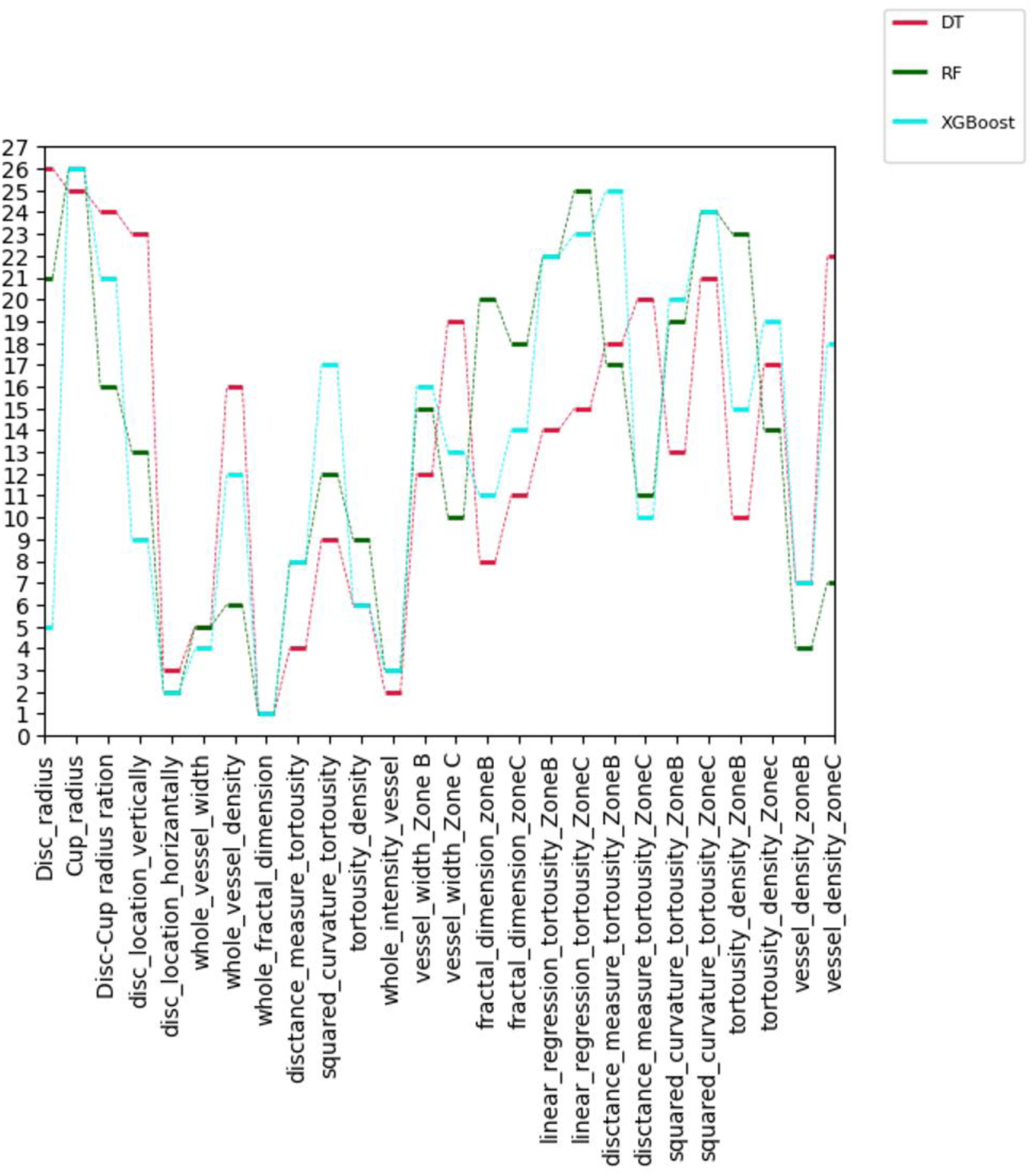
visualization of the feature ranking with RFE method by using three different classifiers, DT, RF and XGBoost, as the estimators. The number 1 on the y-axis indicates the first important feature and so on.

Comparing the results obtained for feature importance by XGBoost classifier, PI method for three machine learning models (Random Forest, Decision Three and XGBoost classifiers) and RFE method was figured out that three feature vectors including location of Disc horizontally, fractal dimension and intensity level of vessels from the whole image were the most important and valuable features for HC and MS classification.

Then, to visualize these three important features in two dimensions, we used t-distributed stochastic neighbor embedding (t-SNE) known as an unsupervised non-linear dimensionally reduction technique for data exploration to find patterns in lower-dimensional spaces [34]. It works by creating a probability distribution of feature vectors and finding a similar distribution for them as points on the map, meaning that objects that are close to each other in the feature space are more likely to be close to each other on the map. Figure 12 represents the high discriminative capacity of these three important features with t-SNE.

**Figure 12.**
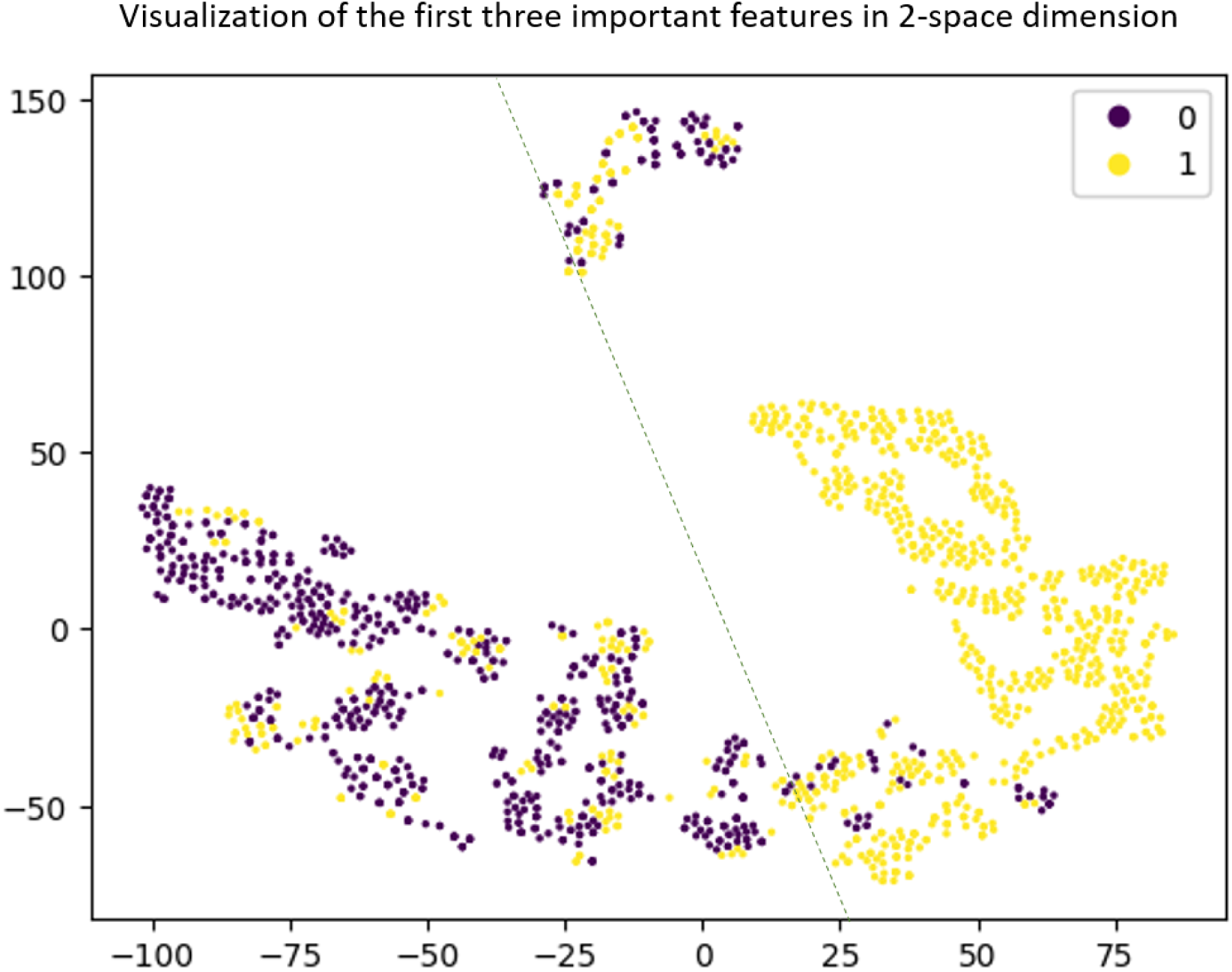
visualization of three important features in 2-space dimension using t-SNE. Label 0: HC images, label 1: MS

These three important features were resulted in a mean ACC of 0.81 (SP=0.82, SE=0.78, F1=0.80, AUROC=0.85, AUPRC=0.87) when testing by the designed XGBoost classifier on validation dataset. This can ensure that these three important features successfully classified the SLO images in two classes, MS and HC, with a performance metric close to what had been resulted from testing on all features.

Evaluating these three important features on age-gender matching test dataset also resulted in a mean ACC of 0.70 (SP=0.78, SE=0.66, F1=0.71, AUROC=0.81, AUPRC=0.91), close to the performance obtained on the whole feature set while testing on the age-gender matching test dataset.

## 4. Discussion

According to the results obtained from the section 3.3, location of Disc horizontally, fractal dimension and intensity level of vessels from the whole image were the most three important features to classify MS and HC. To evaluate these three valuable features, their values on the MS and HC images correctly classified on the all 5 training folds constructed by -fold cross validation (CV) were analyzed compared Regarding the Disc location, it was seen that this could be appeared toward the left or right side of the HC images (before flipping the left eyes), while in MS images this could be located either on the macula region or on the sides of the image. Figure 13 shows the optic disc location in the SLO images of MS patients and HC people. The first row represents the MS images of test dataset correctly classified on 5 training folds, constructed by CV, while the second row illustrates the HC images of test dataset. The location of optic discs is also shown with a red colored rectangle on each image satisfying what is stated earlier.

**Figure 13.**
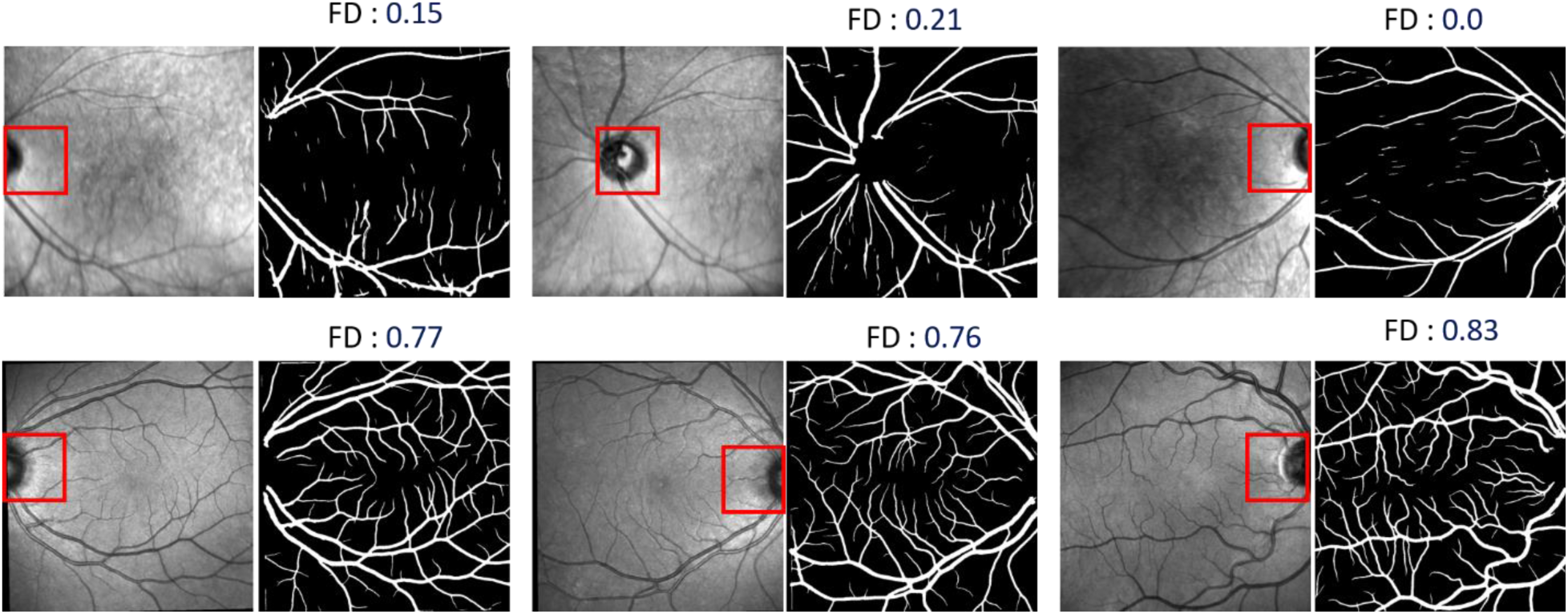
Representation of the first three important features on the images with MS, the first row, and the HC images, the second row. The location of optic discs is shown with a red colored rectangle on each MS and HC image.

Although this feature was selected as the first important feature in our proposed algorithm, we observed that there were two different imaging techniques in capturing the used SLO images, optic disc centered (ODC) and macula centered (MC). So, it did not make sense the location of optic disc could differentiate MS images from HC ones. Therefore, to trust artificial intelligence (AI) results, it is necessary that all the images come from the same imaging protocol.

Respect to fractal dimension (FD), it is a factor measuring how an object details changes at different magnifications. So, Fractal objects shows self-similarity and complexity under a change of length scales [35]. This measurement had been considered as a potential biomarker to recognize some diseases like diabetes and hypertension through description of branching vascular distribution in tow-dimensional space [36]. There are different methods to calculate fractal dimension, including box counting, the mass-radius relation, the two points density-density or pair correlation function method [35]. Box-counting, the most common way to measure fractal dimension, was the method used in this work to determine fractal dimension in the SLO images. It works by overlaying the binary image (the map of segmented vessels) with a grid of boxes of side length ɛ and counting the number of boxes containing a part of vessel tree. This process is repeated under different value of ɛ to obtain more and more fine details of the vascular tree from the covering. Finally, the box-counting dimension can be calculated through following formulate in which *N*(∈) represents the number of boxes containing vascular tree:

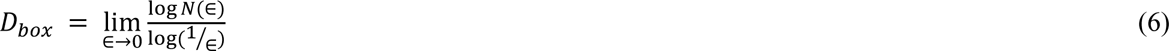

Figure 14 shows the process of calculating FD for an SLO image based on box-counting method. The SLO image and its segmented blood vessel map are shown in the first and second columns, respectively. The third and fourth columns represents what is stated in equation 5.

**Figure 14.**
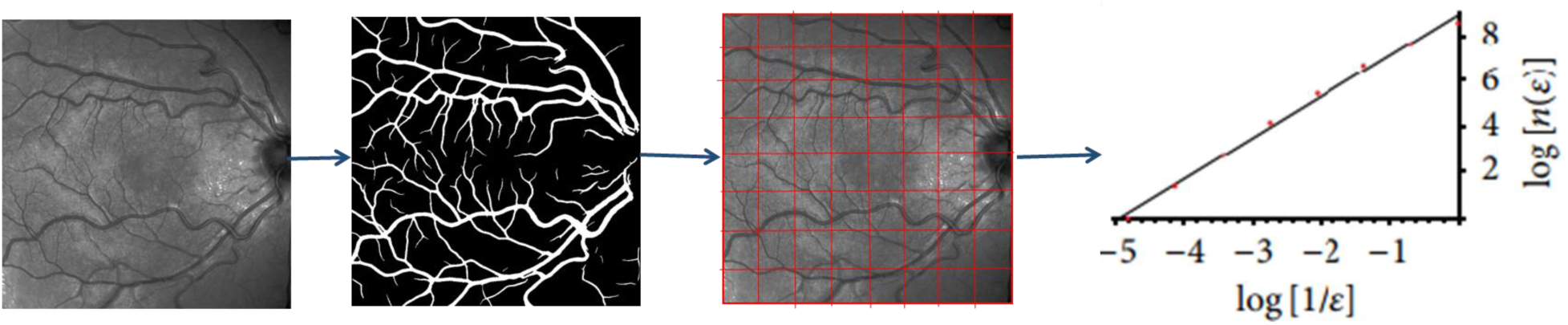
the process of measuring FD for an SLO image.

In figure 13, it was seen that the value of fractal dimension was more in HC images compared to its value on the MS images. It should be noted that the value of FD is normalized by min-max normalization in which the FD values range gets transformed into the range from [0,1) that does not affect the role of this feature. In this figure, it can be also seen that the MS images (the first row) are not self-similar based on box-counting method, while the complexity and similarity of blood vessels in HC images (the second row) can be clearly seen.

And finally regarding to the ratio of intensity of blood vessels to intensity of whole image, it could be seen in some SLO MS images correctly classified as MS, not only there were some white patches in background of the images, but also the images were blurrier than the average scan. In figure 13, the impact of this feature on MS and HC classification is visible. As it was shown, there were some white patches on the SLO images with MS, resulting in lower values for this feature.

It has been previously shown that the blood flow velocity (BFV) of retinal arteries and veins in MS patients is significantly lower than HC people [37]. Decreased blood flow will decrease the content of oxygen in the blood vessels. Blood oxygenation, the measure of oxygen present in arterial or venous blood, is a measure of how much hemoglobin is currently bound to oxygen compared to how much hemoglobin remains unbound. So, the low content of oxygenated hemoglobin in arteries absorb more light and therefore reflect low light [38]. This can be figure out as the main reason for the low values of the ratio of blood vessel intensity to intensity of the whole image in MS patients compared to HC subjects.

Although this study demonstrates the most important and effective features of SLO images for classification of MS and HC subjects, there are several limitations to be addressed, helping to achieve more accurate results related to segmentation and feature extraction. Fist, the train and validation SLO images used for this study were considered of a limited number of samples obtained from a single center. Furthermore, this could lead to improvement of the proposed methods for anatomical segmentation of optic disc, cup and blood vessels, if we have had their ground truth images, which in turn could end up in achieving more accurate results for the feature extraction step. Finally, if calculated, the arteries’ and veins’ features could result in improving the important and valuable feature set extracted from the SLO images for MS and HC classification.

## 5. Conclusion

To conclude, the aim of this work was to calculate the most important, effective and discriminating clinical features from SLO images to classify MS and HC subjects. To achieve this purpose, we decided to compute a set of manual features of some retinal structures, including optic disc, cup and blood vessels, to get a better understanding of the features that could be relevant for the classification of MS and HC patients. Therefore, the optic disc, cup and blood vessels had to be first segmented from the SLO images and then used for feature computation. Following this, we took advantage of three different simple machine learning models, SVM, RF and XGBoost, to evaluate the obtained feature set and then computed the importance of each feature by using three different methods. The disc location, fractal dimension and intensity variations of vessels were selected as the most important clinical features. As the used SLO images had been captured with 2 different imaging protocols named ODC and MC, optic disc location could not be trusted as a valuable feature. Fractal dimension and intensity of vessels, two other important features, were also confirmed by one expert, showing the consistency of these effective features for MS and HC classification.

## Data Availability

data is not available

