## Supplementary for "Analyzing morphological alternations of vessels in multiple Sclerosis using SLO images of the eyes"

#### A brief introduction of the algorithm used for calculating the boundary of optic disc candidate located on right side of the SLO image:

To obtain the boundary of optic disc candidate located on the right side of the image, the used blob algorithm failed to correctly detect it. To address this problem, we used the information obtained from the blob algorithm to estimate the optic disc candidate boundary.

Figure 1 represents a SLO image with the optic disc candidate located on the right side of the image and the image containing the optic disc boundary obtained from blob algorithm.

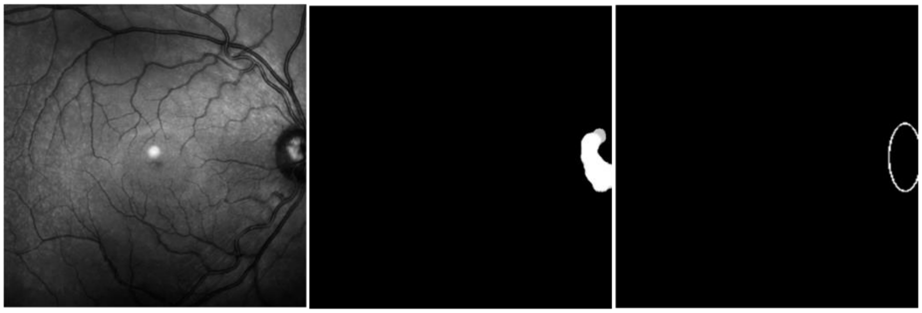

Figure 1. visualization of a SLO image, the left image, its optic disc candidate, the middle image and the result obtained from blob algorithm, the right image.

Figure 2 displays the algorithm we used in such cases that blob algorithm alone was not able to measure the boundary of optic disc candidates. As the first step, we needed to select three points with the greatest distance from each other on the boundary of the ellipse obtained from blob algorithm. In figure 2, these three points satisfying the criterion are shown as three red colored points named A, B, C. At the second step, after calculating the line segments AB and BC, shown as yellow and blue colored line segments respectively, we computed the perpendicular bisector of each of these line segments which are shown with two green colored line segments in figure 2. The point resulting from the intersection of two calculated perpendiculars was considered as the center of the optic disc candidate, named with D in this figure. Finally, the boundary of the optic disc candidate was measured with the help of blob algorithm using point D and length of the line segment BD, as the center and radius of the optic disc candidate, respectively, shown with the green circle colored on the SLO image in figure 2.

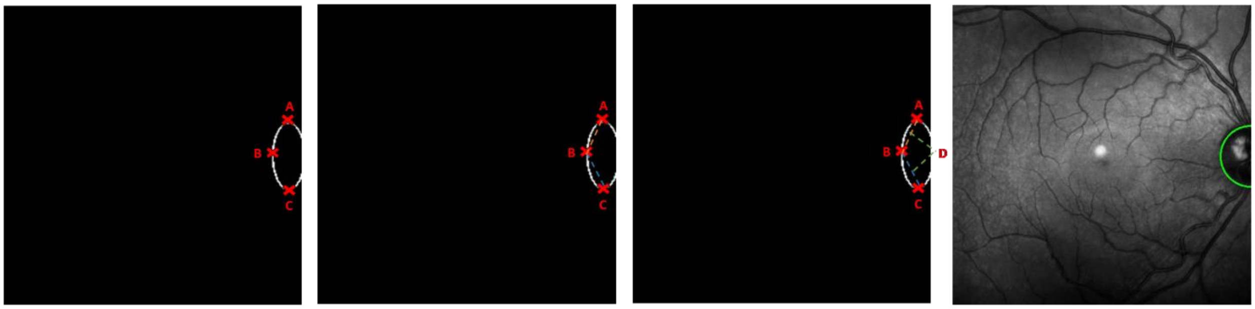


Figure 2. Visualization of the algorithm used for estimating the boundary of the optic disc candidate located on the right side of the SLO image. Representation of three points A, B,and C with the greatest distance from each other, the left image, two line segments AB and BC and their perpendiculars and finally the boundary of the optic disc candidate with the green colored circle, the right image.

### A brief introduction of the reference image used in vessel segmentation process:

To segment blood vessels from the SLO images in our dataset, we first needed to manipulate the intensity histogram of the SLO images to address the variations of intensity values which could lead to the detection of false vessel pixels. For this purpose, we used a histogram matching algorithm in which the histogram intensity of a reference image was used to unify the contrast level of other SLO images. Figure 3 represent a SLO image with intensity variation, a reference image and finally the modified contrast level of the SLO image according to the reference image. The reference image was chosen in such way that not only its vascular tree satisfied a high value of intensity recognizable form the background, but also the intensity variation of the background pixels was low and negligible.

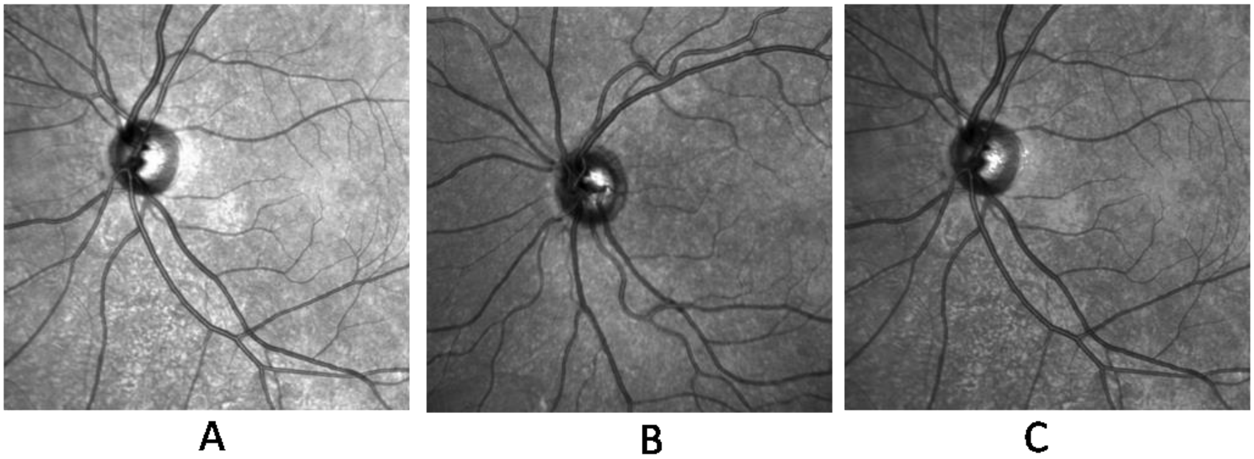


Figure 3. A visualization of the SLO image with intensity variations in its background, image A, the reference image used for matching histogram algorithm, image B, and the SLO image with matched contrast level according to the reference image, image C.

### A brief introduction of zone B and zone C from the center of optic disc:

In current work, there were 2 standard areas centered on the optic disc of the SLO images used for vascular feature measurements, named as zone B and zone C. Figure 4 illustrate these two areas on a retinal image in which zone B defines a circumferential region 1 to 1.5 optic disc diameter from the center of the optic disc and zone c is related to a circumferential region 1 to 2.5 optic disc diameter form the optic disc center.

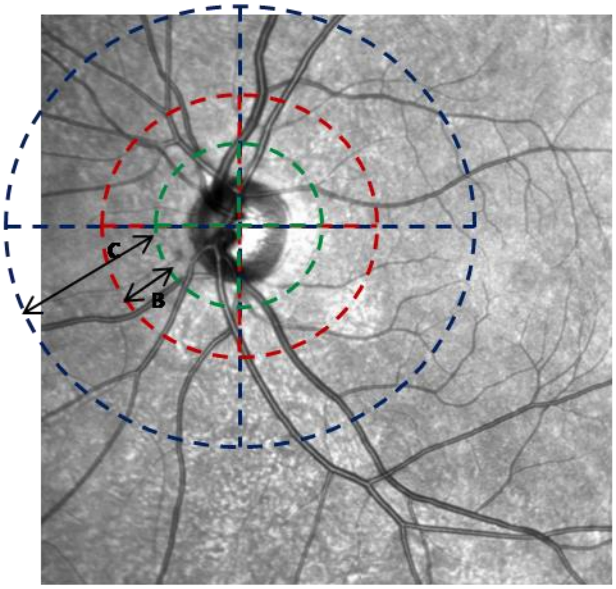


Figure 4. SLO photograph indicating ZONE B and zone C.
